# Efficacy of Pulmoary exercise administered in Supine, 45°, or 90° long sitting position on pulmonary parameters of patients with COPD: A Study Protocol

**DOI:** 10.1101/2022.01.30.22270106

**Authors:** Eniola Awolola, Sonill Maharaj, Oluwafemi Ojo, Olufunke Adeyeye

## Abstract

**Background:** Pulmonary rehabilitation is a program commonly structured for the rehabilitation of chronic lung diseases like COPD, chronic bronchitis, and emphysema. It aims to increase the patients’ quality of life by decreasing the symptoms frequently experienced from respiratory diseases.

**Methods:** Forty-two (42) patients aged 40 and above with Chronic Obstructive Pulmonary Disorders will be assessed for eligibility for the study. The study design will be a double-blinded, randomized control trial with three intervention groups and three parallel placebo control groups.

ACBT, Autogenic drainage, and Reciprocal pulley exercise will be administered as an intervention in various body positions for 5 minutes, respectively. Ten continuous outcome variables at a different point with a minimum repeated measurement of six per subject will be utilized as an outcome measure. Baseline Pulmonary Function Test (PFT) will be assessed on entry into the study and after every procedure. MRC breathlessness scale, six minutes’ walk (6MWT) test, and CAT assessment test will be assessed at every two (2) weeks of the study. Data will be analyzed using descriptive and inferential statistics of repeated ANOVA; P<0.05.

**Discussion:** The study outcome will determine pulmonary rehabilitation’s efficacy in various body positions on selected pulmonary parameters of COPD patients and identify the most productive approach among upper limb exercises, autogenic drainage, or ACBT in different fundamental body position

**Trial Registration:** www.pactr.org: PACTR202005890624077

## Background

Chronic obstructive pulmonary disease (COPD) is a group of airway diseases commonly associated with emphysema, chronic bronchitis, or both. Emphysema and bronchitis can lead to airway obstruction following the destruction of air sacs in emphysema and mucus build-up from inflammation in the case of bronchitis (1, 2). According to <u>Similowski, Agusti</u> (3), COPD is characterized by progressive, partially reversible airflow obstruction with significant extrapulmonary (systemic) manifestations, lung hyperinflation, and comorbid conditions that may add to the severity of the disease in individual patients.

Pulmonary rehabilitation (PR) is a multidisciplinary, comprehensive intervention that is evidence-based for patients with chronic respiratory diseases who present with symptoms and reduce functional activities of daily living (4-6). PR’s essential components are education, self-management training, nutrition, and psychological support, while Exercise training is the cornerstone of the PR (6, 7). PR has demonstrated an increased exercise tolerance, lower dyspnea perception, and improved quality of life in COPD patients. Responses to PR differ in patients, and no improvement can be achieved in some patients (6, 8).

According to (9, 10), the presence and number of comorbidities may decrease PR’s positive effects on perceived breathlessness, functional exercise capacity, and quality of life.

Body posture has long been identified as an essential factor impacting lung volumes (11). Also, body positions are clinically significant in a healthy population during treatment, resuscitation, everyday activities, and surgical procedures (12). Although the effective patient positioning may be associated with marked improvement in PaO2, some positions may deteriorate V/Q matching (13).

Even though randomized controlled trials have indicated the efficacy of upper limb exercises, autogenic drainage, and ACBT in COPD patients’ rehabilitation, none has considered the pulmonary rehabilitation impact varied across different fundamental body positions on patients’ pulmonary parameters. This study is designed to evaluate a 12-week efficacy of pulmonary exercises administered in Supine, 45° long sitting and 90° long sitting program on FVC, FEV1, and FEV1/FVC of patient’s with COPD.

## Statement of the problem

In managing COPD, lack of awareness, knowledge, and often delay in recognizing disease are reasons why health care providers may incorrectly diagnose or manage the condition (14). Clinical diagnosis of COPD is considered in any patient with dyspnea, chronic cough or sputum production, or a history of exposure to risk factors for the disease, which is often confirmed by spirometry (15).

According to <u>Solomen</u> (16), persons with chronic cough symptoms, sputum production, shortness of breath, or wheezing, especially those with prolonged exposure to disease risk factors, should be considered evaluated for COPD. The standard features are shortened Inspiratory to expiratory (I: E) ratio, pursed-lip breathing (PLB), recruitment of accessory muscles during respiration, jugular venous distension, labored breathing signs, pulsus paradoxus, barrel□shaped chest, peripheral edema, dyspnea, muscle wasting, and, restricted chest expansion, diminished breath sound, early inspiratory crackles, a loud pulmonic component in the second heart sound and rhonchi during expiration (17, 18).

A pulmonary function test is a tool to assess airflow obstruction. The forced expiratory volume in 1 s (FEV1)/forced vital capacity ratio of patients is reduced, and FEV1 is reduced in COPD (19). A reversibility test differentiates COPD from Asthma; patients with COPD do not show reversibility in airflow obstruction (19). Severity staging of COPD is important as an outcome tool and as well as for treatment. The GOLD guidelines classifies COPD into mild (FEV1 ≥ 80% predicted), moderate (50% ≤ FEV1 < 80%), severe (30% ≤ FEV1 <50%), and very severe (FEV1 <30%) disease (19).

Pulmonary rehabilitation (PR) is an evidence-based, multifaceted, and comprehensive intervention for patients with chronic respiratory diseases who presents with symptoms and often have decreased daily life activities (4, 6, 20). Education, self-management training, nutrition, and psychological support are essential PR components; exercise training is the cornerstone of the PR (6, 7). PR has increased exercise tolerance, lower dyspnea perception, and improved quality of life in COPD patients. The patients’ responses to PR differ, and some can not improve (6) (8).

According to <u>Hornikx, Van Remoortel</u> (9), (10), other factors like comorbidities may decrease PR’s positive effects on symptoms frequently experienced by COPD patients.

Even though randomized controlled trials have indicated the efficacy of upper limb exercises, autogenic drainage, and ACBT on COPD patients, none has considered the PR impact varied across different fundamental body positions on the pulmonary parameters of COPD patients. This study is aimed to evaluate a three months efficacy of pulmonary exercises administered in Supine, 45° long sitting and 90° long sitting program on MRC dyspnea scale, Six minutes’ walk, CAT score, HR, RR, SBP, DBP,FVC, FEV1, and FEV1/FVC of patients with COPD. This study is structured to find the solution to the listed questions.

1. What will be the effect of pulmonary rehabilitation applied in Supine, 45° or 90° long sitting on MRC dyspnea scale, Six minutes’ walk, CAT score, HR, RR, SBP, DBP, FVC, FEV1, and FEV1/FVC of patients with COPD over three months?
2. Will PR applied in Supine, 45° or 90° long sitting affect MRC dyspnea scale, Six-minute walk, CAT score, and Cardiovascular parameters of patients with COPD over three months?
3. Will PR applied in Supine, 45° or 90° long sitting affect MRC dyspnea scale, Six minutes’ walk, CAT score, and Pulmonary parameters of patients with COPD over three months?

### Aim of the study

The overall aim of the study is to determine the effect of PR applied in Supine, 45° or 90° long sitting on MRC dyspnea scale, six minutes’ walk, CAT score, HR, RR, SBP, DBPFVC, FEV1, and FEV1/FVC of patients with COPD patients attending respiratory clinic of Lagos State University Teaching hospital, Ikeja. Lagos.

### Specific Objectives

To determine

i. The effect of PR in 3 fundamental body positions on the MRC dyspnea score of patients with COPD attending the respiratory clinic.
ii. PR’s impact in 3 fundamental body positions on 6 minutes’ walk test score of patients with COPD attending the respiratory clinic.

### Hypotheses

#### Main Hypothesis

##### Ho

Pulmonary Rehabilitation (PR) applied in Supine, 45° or 90° long sitting will have no significant effect on MRC dyspnea scale, Six minutes’ walk, CAT score, HR, RR, SBP, DBP, FVC, FEV1, and FEV1/FVC of patients with COPD over three months.

##### Ha

Pulmonary Rehabilitation (PR) applied in Supine, 45° or 90° long sitting will have a significant effect on MRC dyspnea scale, Six minutes’ walk, CAT score, HR, RR, SBP, DBP, FVC, FEV1, and FEV1/FVC of patients with COPD over three months.

#### Sub Hypotheses

i. There will be no significant difference in the MRC dyspnea scale, Six minutes’ walk, CAT score before and after pulmonary rehabilitation administration in Supine, 45° long sitting, and 90° long sitting study group.
ii. There will be no significant difference in the MRC dyspnea scale, Six minutes’ walk, CAT score before and after pulmonary rehabilitation administration in Supine, 45° long sitting, and 90° long sitting of the control group.
iii. There will be no significant difference in the MRC dyspnea scale, six minutes’ walk, CAT score, and Cardiopulmonary parameters (SBP, DBP, HR, RR, FEV1, FVC, AND FEV1/FVC) before and after administration of pulmonary rehabilitation in Supine, 45° long sitting and 90° long sitting of the two groups at baseline, 2nd. 4th, 6th, 8th, 10th, and 12th week of intervention.

### Delimitation

This study will be delimited to 42 patients with COPD attending Lagos State University Teaching Hospital’s respiratory clinic, Ikeja, Lagos.

### Significance of the study

It is expected that the outcome of this study will establish the relationship between Pulmonary rehabilitation and body positions on the MRC dyspnea scale, CAT scores 6 minutes’ walk and Cardiopulmonary variables of patients living with COPD.

The outcome of this study will provide empirical evidence on the Physiotherapy management of COPD.

## METHOD

### Study Design

The study is a 12-week randomized control trial. The study will entail six (6) groups, three intervention groups, and three placebo control groups.

### Participants

This study will consist of male and female adult COPD patients aged 40 years and above attending Lagos State University Teaching Hospital (LASUTH), Ikeja, Lagos, Nigeria, West African.

*The inclusion criteria involve patients with a COPD history of about or more than six months of duration attending the respiratory clinic of LASUTH*.

*The exclusion criteria involve patients with COPD on a cardiac pacemaker, supplemental oxygen therapy, Asthma, cardiac conditions, and patients with psychological impairments*. Participants who meet the inclusion and exclusion criteria will be required to read and sign an informed consent approval for this study by the appropriate ethical institutional review board.

### Setting

Patients with COPD attending the respiratory clinic of Lagos State University Teaching Hospital Ikeja, Lagos State, Southwest Nigeria, would be recruited for the study.

The hospital is a tertiary health facility within the state.

### Sample size

Pulmonary function test is the primary outcome of interest for the study and the expected clinically relevant difference for pulmonary rehabilitation in various body positions using LLN and GLI reference equation proposed by Quanjer PH (21). Therefore, the sample size (N) will be determined using G-Power statistics software. The power is selected at 95% =0.95, confidence level at 5% =0.05 and effect size of 0.35. We intend to investigate the main effects, within and between interactions for two factors, namely:

Factor A – Therapy with two levels (No Therapy and Therapy)

Factor B – Positioning of patients with three levels (Supine, 45 and 90 long sitting), nested within factor A

This gives 2×3 = 6 groups of subjects from which repeated measurements are to be obtained. We are also planning to measure ten continuous outcomes at different time points. The minimum repeated measurements are expected to be 6 per subject for some of the outcomes and a maximum of 36 per subject. Suppose a minimum of 6 repeated measures is to be obtained from each subject. In that case, it is estimated that a minimum total sample size of 42 patients is required to detect an effect size v = 0.35 with 95% confidence (5% Type I) and having similar studies also expected to produce similar results about 80% of the time (Power of test 80%). In the case that during the time of the data collection, the hospital experiences a heavy burden of COPD patients or resources permit, there is an allowance to increase the total sample size to at most 59 subjects with a corresponding increase in the power of the test to about 95% and still detecting effect sizes between 0.3 and 0.4 (See Figure 1). This translates to the possibility of recruiting between 7 and 10 patients per group. Hence, the sample size estimates suggest that recruiting more than ten patients per group for this kind of study will be a waste of resources.

**Fig 1.**
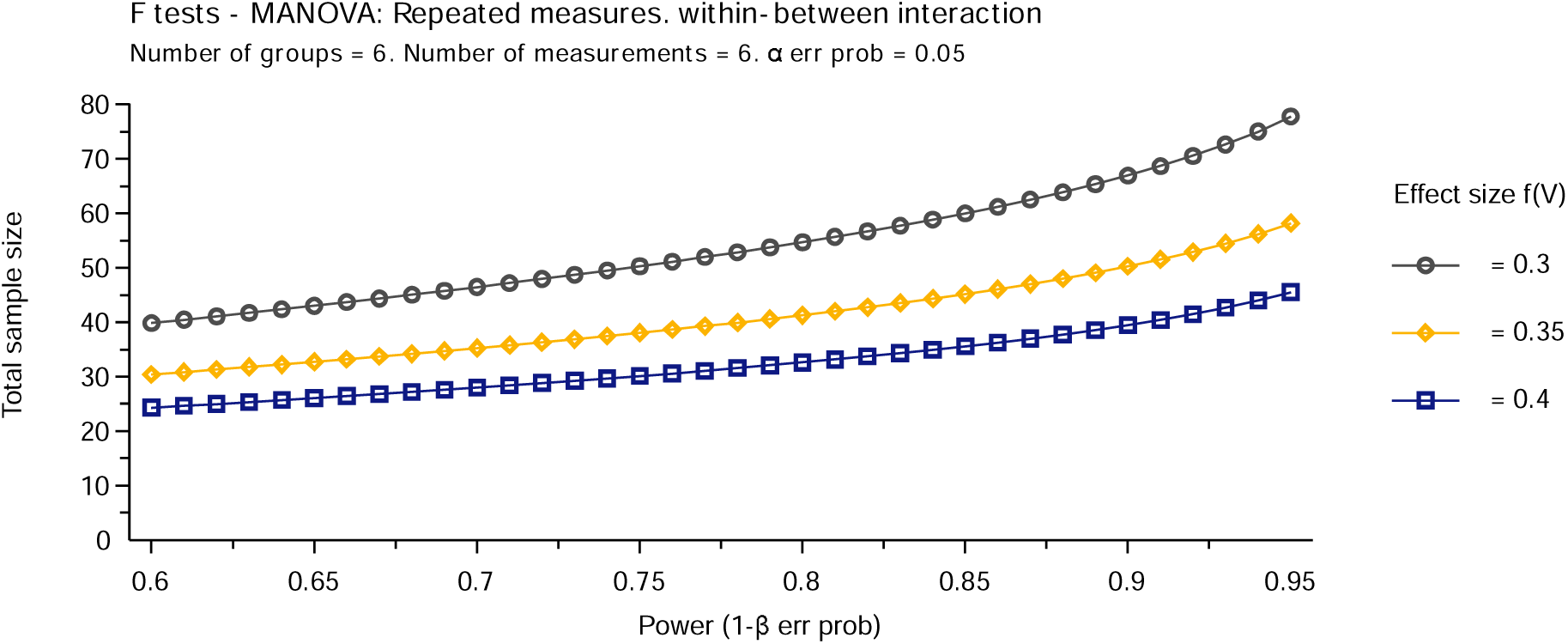
Sample size calculation.

### Randomization and blinding

The contact numbers of participants will be randomly extracted from the respiratory patients’ database attending Lagos State University Teaching Hospital Ikeja’s respiratory clinic. A bulk Text message captioned “Invitation to a study on COPD will be circulated using the Luxury bulk SMS platform. Respondents will be assessed for eligibility, and those that meet the inclusion and exclusion criteria will participate in the study.

Participants will be randomly selected by simple randomization using a computer software program (Suresh, 2011). We will use the software program www.randomization.com to allocate participants into study group A and control group B. Group A will be further assigned to subgroup x, y, z, and group B will be assigned to subgroup e, f, g. Subgroup x and e connote supine position, y and f connote 45°long sitting, while z and g represent 90° long sitting position, respectively (Figure 2).

**Fig 2.**
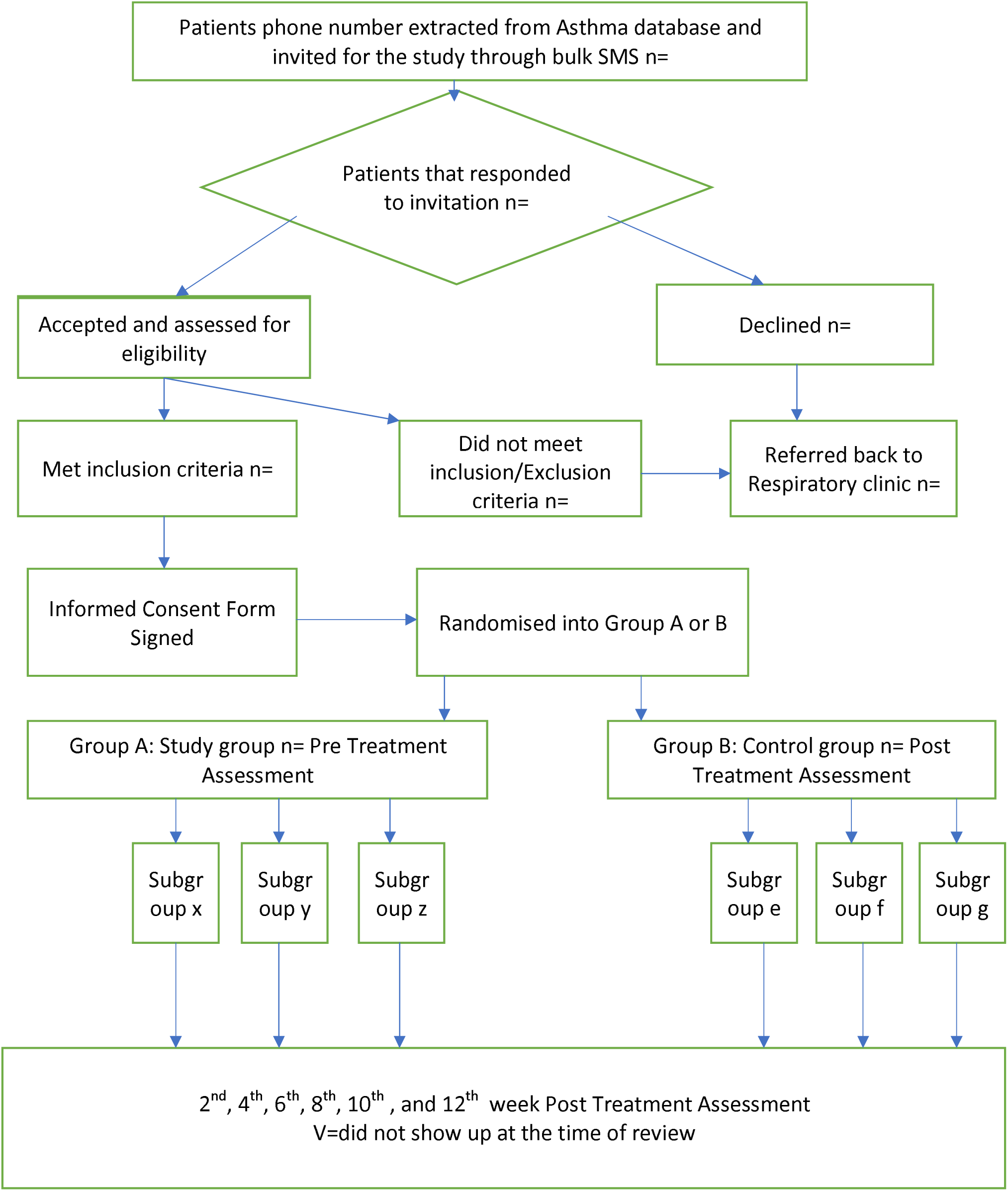
Recruitment and Randomization of Participants.

### Intervention

Participants will be briefed about the nature of the study, the effect and benefit of the study. They will be encouraged to clarify issues regarding the study; if any, written informed consent will be obtained from participants before the study’s commencement. Participants will be randomly assigned into two groups: Study (Group A) and Control group (Group B). Six minutes’ walk test, MRC dyspnea scale, and CAT score will be measured and recorded before intervention in both groups. Reassessment will be done after the 2nd, 4th,6th, 8th, 10th, and 12th week of the study intervention.

The study group (Group A) will receive reciprocal pulley exercise, Autogenic drainage, and ACBT, with each procedure not exceeding five minutes. If the patient presents any sign of distress during treatment, the treatment will be immediately discontinued, and the patient will be placed under close observation. If the situation persists, the patient will be referred to the respiratory unit for further reevaluation. So long as the subject experiences no distress, the intervention will last a total of twenty (20) minutes with an average rest time of 90 seconds between procedures.

### Autogenic Drainage

The subjects will be asked to blow their nose and clear the throat before commencing the procedure with the subject in the test position (Supine, 45° long sitting or 90° long sitting). The technique will be explained to the participants as follows; the technique involves three phases: the unsticking phase, collecting phase, and evacuating phase. The first stage involved the subject taking a deep breath, then breathing out completely. After this, the subjects will be required to take a small breath and breathe out completely. When the subject feels the secretions’ movement, the subject will move to the collecting phase. The subject will be required to take a deeper breath, and the expiration should not go very deep into the expiratory reserve volume. Once the secretion is felt moving to more central airways, the subjects will be asked to take very deep breathes to reach the inspiratory reserve volume and enter the evacuating phase. As the subjects feel that the secretions have gone high in the central airways, they will be required to cough out the sputum into the sputum tray beside the treatment bed. The subjects will be asked to control the cough during the cycle of breathing exercise (22). The cycle will be repeated for 5 minutes

### ACBT

With subjects in the fundamental starting position (Supine, 45° long sitting or 90° long sitting). ACBT will be explained to the participants as follows; ACBT involves three stages: breathing control, deep breathing exercise, and Forced Expiratory Techniques (FET). The cycle consists of a sequence of breathing control. The subject will be required to do quiet, relaxed abdominal breathing, deep breathing exercises in which the subject will be required to take 3 or 4 deep breaths to expand the thoracic region. The first two stages will be repeated five to six times before reaching the third stage, which is the FET in which the subject will perform 1 or 2 huffs, from low lung volume followed by huff from high lung volume and relaxed abdominal breathing (22). The entire technique will be continued for 5 minutes.

#### Control group

The control group (Group B), in addition to the baseline pulmonary function test, will also receive counseling on COPD and enjoy free musculoskeletal assessment. They will be divided into three subgroups and labeled e, f, and g, placed in Supine, 45 degrees, and 90 degrees long sitting. The participants will be required to maintain the position for 20 minutes. If the subject is experiencing respiratory difficulty, the procedure will be discontinued.

### Outcomes

#### Six-minute walk Test

According to ATS (23) six-minute walk test (6MWT), a practical, simple test requires a 100-ft hallway with no exercise equipment or advanced training. It measures the distance that a patient can quickly walk on a flat, firm surface in 6 minutes (the 6MWD). It evaluates the global and integrated responses of all the systems involved during exercise, including the pulmonary and cardiovascular systems, systemic circulation, peripheral circulation, blood, neuromuscular units, and muscle metabolism.

#### MRC Dyspnea scale

The MRC breathlessness scale quantifies the disability associated with breathlessness by identifying if breathlessness occurs when it should not (Grades 1 and 2) or by quantifying the associated exercise limitation (Grades 3–5) (24).

According to Mahler and Wells (25), there is up to 98% agreement between observers recording MRC breathlessness scores. The score correlates well with the results of other breathlessness scales, lung function measurements, and direct measures of disability such as walking distance (23).

The MRC breathlessness scale is commonly used to describe patient cohorts and stratify them for pulmonary rehabilitation interventions, predict survival and use as complementary to FEV1 in describing disability in those with COPD (26-28). It has demonstrated its worth in 50 years of use (25).

#### CAT Score

According to P.W. Jones (29), the CAT was created using COPD patients’ input, developed using modern questionnaire methodology, psychometric analysis, and item response theory using Rasch analysis to identify items with the best fit to form a unidimensional instrument. It is a self-administered questionnaire consists of eight items assessing various manifestations of COPD (cough, sputum, dyspnea, chest tightness) aiming to provide a simple quantifiable measure of HRQoL.

CAT scores of 0–40 denote the severity of COPD; the higher the score, the more significant the impact of COPD on a patient’s life, its intra-class correlation coefficient=0.8, its internal consistency Cronbach’s α=0.88. The instrument has a high correlation with SGRQ (r=0.84) across 7 European countries (29).

## SPIROMETER

Koko SX 1000 Standalone Version 7 Pneumotach, a portable lightweight and comprehensive diagnostic tool will conduct pulmonary function tests.

The Koko Legend II spirometer has a built-in thermal printer and a touch screen display. It can perform FVC, Pre vs. Post, and SVC tests. Test data and patient information is stored directly on an internal SD card that can be replaced and re-used if necessary. All the stored data can be downloaded via a USB cable onto a PC for backup or storage. This device supports daily calibration checks, complies with ATS-ERS 2005, has several Predicted Authors, and includes GLI-2012.

Daily calibration of the device will be conducted using a 3L syringe.

The participant’s condition can be shown by the ratio of the measured value and the predicted value. Flow rate-volume chart, volume-time chart display, data memory, delete, upload and review, trend chart display, scaling (Calibration), information prompts when volume or flow goes beyond the limits are features available in the device.

### Data analysis

We will use the Statistical Package for Social Sciences (SPSS Inc, Chicago, II) version 26.0 for the Windows package program to analyze data. We will use descriptive statistics of mean, standard deviation, frequency, and percentage to summarize the results. Bar charts, pie charts, and histograms will be utilized for pictorial illustration.

A multilevel analysis of variance (MANOVA) will be used to compare the outcome variables [selected cardiovascular variables (RR, HR, SBP, and DBP), body position (Supine position, 45 degrees long sitting and 90 degrees long sitting), pulmonary function variables (FEV1, FVC, FEV1/FVC), MRC dyspnea scale, Six minutes’ walk and CAT score among each group, while dependent t-test will be used to compare the pre and post-intervention results. An Independent t-test will be used to compare the outcome variable across the two groups. The level of significance will be at p0.05

### Harms

All interventions in this study can only be performed within the patient’s tolerance; hence, the research does not pose any potential danger. Occasionally participants may experience mild discomfort from the exercise, including temporary muscle soreness, sweating, and dizziness. Necessary care will be in place to prevent the occurrence of an adverse event. During the trial or after an intervention, in case of a report of serious adverse events like comorbidities, injuries, persistent excruciating pain, dizzy spells, headache, etc., we would consider unblinding the participant to the intervention for his/her safety. Additionally, the participants will be instructed to report any adverse events to the PI or the physiotherapist supervising their group. For adequate supervision and safety, we will limit the number of participants per group to a maximum of 3. Arrangements have been made with the hospital’s Accident and Emergency units, where we will conduct the research to provide a standby medical team. However, the University of KwaZulu-Natal insurance scheme on clinical trials has fully covered the participants in this type of study.

## Discussion

Even with a sizable number of randomized control trials on the effectiveness of pulmonary rehabilitation in managing COPD, information on PR’s effectiveness in different body positions is lacking.

According to Jibril Mohammed (30), at the Aminu Kano Teaching Hospital, Kano, on the effect of different body positioning on lung function variables among patients with bronchial Asthma, FEV1 and FVC increased in the standing position compared to sitting and supine position among 20 participants.

On the contrary, (31) in a systematic review further reported no statistical difference in patients’ pulmonary parameters with COPD in sitting vs. Standing position.

We expect that this study’s findings will give a clearer understanding of pulmonary exercise’s effect on selected fundamental body positions of patients with COPD and serve as a guideline for pulmonary rehabilitation in managing COPD patients.

## Access to Protocol

https://pactr.samrc.ac.za/Researcher/ManageTrials.aspx The protocol was registered on 1st of May 2020 with identifier number PACTR202005890624077 and PACTR is the trial organization.

## Data Availability

The datasets used during the current study are available from the corresponding author on request. The findings from the study would be made available to participating researchers as required by law. All data obtained are stored in a retrievable file system located at the department of Physiotherapy, university of Kwazulu-Natal, South Africa.

## Acknowledgments

We would like to appreciate the numerous support of physiotherapists and medical doctors in Lagos State University Teaching Hospital Ikeja, Lagos, during the early phase of the development of this protocol, especially during the process of fact-finding on the availability of patients for the proposed study from the respiratory unit of the department of Medicine of Lagos State University Teaching hospital, Ikeja, Lagos. Our profound appreciation goes to Dr. Olufunke Adeyeye and Mr. Adewunmi Olalekan of the hospital proposed for the study, for their professional advice in writing and reviewing this manuscript, also Miss Kemi and Miss Laide for their support in the Pulmonary function test, and Miss Amodeni Ayomopewa for her editorial input.

## Ethical Considerations and consent to participate

The Biomedical Research Ethics Committee of the University of KwaZulu Natal (South Africa) has approved this study with approval number BREC/00001883/2020. The Human Research Ethics Committee of Lagos State University Teaching Hospital, Ikeja, Lagos, Nigeria, West Africa, where we will conduct the investigation, has been approved with approval number LREC/06/10/1428. The study has been registered with ClinicalTrial.gov with the following registration number: PACTR202005890624077. All participants will be allowed to read and sign the informed consent before the commencement of the study. A third party who is independent of the survey shall present the informed consent. The consent form is designed by the Biomedical Research Ethics Committee of the University of KwaZulu-Natal (BREC) according to the WMA Helsinki declaration and good clinical practice (GCP).

In the event of the need to modify or amend the protocol, especially inclusion or exclusion criteria of the study, the PI will communicate in writing to the RECs.

## Consent to publication

Not Applicable

## Availability of data and materials

The datasets used during the current study are available from the corresponding author on request. The findings from the study would be made available to participating researchers as required by law.

## Competing interests

The authors declared that they have no competing interests.

## Funding

Awolola Eniola Oladejo funds the study. The institution has no interest or role in the study’s design, writing the manuscript, collecting data, and analyzing data. No external funding is received from any source for the study.

## Abbreviations

ANOVA: Analysis of Variance
COPD: Chronic Obstructive Pulmonary Disease
PEFR: Peak expiratory flow rate
FVC: Forced Vital Capacity
FEV1: Forced expiratory volume in 1 second
HR: Heart rate
RR: Respiratory rate
SBP: Systolic Blood pressure
DBP: Diastolic blood pressure
MRC: Medical Research Council
CAT: COPD assessment Test
6MWT: Six minutes walk test
6MWD: Six minutes walk distance
PFT: Pulmonary Function Test
GLI: Global Lung Initiative
LLN: Lower Level of Normal
FRC: Functional residual Capacity
LASUTH: Lagos State University Teaching Hospital
MANOVA: Multiple Analyses of Variance
MS: Microsoft
RCT: Randomized Control Trial
ROM: Range of Motion
SPSS: Statistical Package for Social Sciences
UKZN: University of Kwazulu-Natal

## Authors’ Contributions

AOE developed the study’s idea; AOE and MSS (developed the title, and all contributed to the study design. All authors were involved in designing the Pulmonary rehabilitation procedure and selecting outcome measures for the study. AOE was responsible for drafting the initial manuscript. OO helped with editing and critical review of the manuscript to add value to its intellectual content. All authors read, critically revised, and approved the final version of the manuscript.

## Notes

### Competing Interest Statement

The authors have declared no competing interest.

### Clinical Trial

PACTR202005890624077

### Clinical Protocols

https://www.pactr.org

### Funding Statement

This study did not receive any funding

### Author Declarations

Human Research Ethics Committee of Lagos State University Teaching Hospital, Ikeja, Lagos, Nigeria, West Africa, approved the study with approval approval number LREC/06/10/1428

